# Per vaginal bleeding-an important but ignored feature of dengue

**DOI:** 10.1101/2024.12.23.24319534

**Authors:** Ananda Wijewickrama, Heshan Kuruppu, Damayanthi Idampitiya, Rivindu Wickramanayake, Anagi Kottahachchi, Jeewantha Jayamali, Padukkage Harshani Chathurangika, Nushara Senatilleke, Navanjana Warnakulasuriya, Chandima Jeewandara, Gathsaurie Neelika Malavige

**Author notes:** Corresponding author information: Prof. Neelika Malavige DPhil (Oxon), FRCP (Lond), FRCPath (UK), Allergy Immunology and Cell Biology Unit, Department of Immunology and Molecular Medicine, Faculty of Medical Sciences, University of Sri Jayawardanapura, Sri Lanka.

## Abstract

**Background:** Elderly individuals, those with comorbidities and pregnant women are at a higher risk of developing severe dengue and succumbing to their illness. However, an increased incidence of severe dengue and fatalities are seen in females of the reproductive age. As per vaginal (PV) bleeding is an important complication that has not been well characterized, we sought to determine the frequency, complications and disease outcomes in women who develop PV bleeding.

**Methodology/Principal findings:** 288 adult female patients were recruited from the National Institute of Infectious Diseases Sri Lanka. All clinical features and laboratory investigations were recorded throughout the duration of hospital admission along with treatment received.

28/288 (9.72%) reported PV bleeding, with 12/28 (42.85%) developing dengue haemorrhagic fever (DHF) compared to 65/260 (25%) of those who did not have bleeding.

Women who developed PV bleeding were more likely to have developed DHF (OR 2.2, 95% CI 0.98 to 5.1, p=0.06), abdominal pain (OR=2.17, 95% CI = 0.99 to 4.69, p = 0.06), vomiting (OR= 2.0, 95% CI= 0.89 to 4.44, p= 0.10), diarhoea (OR= 4.35, 95% CI = 1.908 to 9.610, p= 0.0004) or evidence of any fluid leakage (OR= 1.98, 95% CI = 0.91 to 4.5, p = 0.11). Although not significant, those who had PV bleeding were more likely to have been given intravenous fluids, blood transfusions and colloids.

**Conclusions:** PV bleeding appears to associate with worse disease outcomes. The possible contribution of PV bleeding to higher incidence of severe dengue and fatality rates observed in many countries, should be further investigated.

**Author summary:** Elderly individuals, those with comorbidities and pregnant women are at a higher risk of developing severe dengue and succumbing to their illness. However, an increased incidence of severe dengue and fatalities are seen in females of the reproductive age. As per vaginal (PV) bleeding is an important complication that has not been well characterized, we investigated the complications and disease outcomes in women who developed PV bleeding, by studying clinical and laboratory characteristics of 288 women with acute dengue. 28 (9.72%) reported PV bleeding, with 42.85% developing dengue haemorrhagic fever (DHF). Women who developed PV bleeding were more likely to have developed DHF, abdominal pain, vomiting, diarhoea or evidence of any fluid leakage. They were more likely to have been given intravenous fluids, blood transfusions and colloids. Therefore, the possible contribution of PV bleeding to higher incidence of severe dengue and fatality rates in many countries should be further investigated.

## Introduction

Dengue was named as one of the top ten threats to global health by the World Health Organization in 2019 [1]. Since then the number of cases has increased exponentially with over 10 million cases being reported in the first six months of 2024 [2]. Dengue is also the only infectious disease where the annual mortality rates are rising, with case fatality rates (CFRs) increasing during outbreaks due to overwhelming of health care systems [2]. Apart from the rise in the number of cases, there has been a gradual shift in the age of infection, with many countries reporting most cases among adults [3, 4]. As a result, more infections occur in older individuals, in those with comorbidities and in women in reproductive age group, some who are pregnant. Infection in older individuals, in those with comorbidities and in pregnancy is known to associate with severe dengue with higher risk of fatalities [5–7]. In the absence of a specific treatment and prognostic biomarkers, increase in infections in vulnerable populations leads a larger proportion of patients developing severe dengue, burdening fragile health care systems.

Although most individuals with dengue experience a mild illness, some develop plasma leakage leading to pleural effusions, ascites, shock, organ dysfunction and severe bleeding [8]. Liver dysfunction, shock, extremes of age, pregnancy and the presence of comorbidities such as diabetes have been identified as risk factors for death due to dengue [9–11]. In addition, females have also shown to be at a higher risk of developing severe disease with higher case fatality rates than males [6, 12, 13]. For instance, although 62% of patients who were hospitalized due to dengue in Bangladesh were males, 58% who succumbed to the illness were females [13]. Furthermore, in Brazil, although most deaths were seen in individuals >70 years of age, among 20- to 49-year-olds, more deaths were seen among females compared to males [5]. In Sri Lanka, although the CFRs due to dengue has markedly reduced over the years (currently 0.05%), in 2024, 19/22 (86.4%) of those who succumbed to the illness were females [14]. 13/19 (68.4%) deaths in women were in those between 15 to 45 years of age [14].

The reasons for a higher incidence of severe dengue and higher mortality among women of the reproductive age groups are not known. These could be due to late presentation to a health care facility due to differences in health seeking behavior among men and women, women presenting to hospital late due to carer responsibilities or because women develop complications due to dengue, more frequently than males. Although gastrointestinal bleeding manifestations are less frequent among dengue patients in Sri Lanka in recent years, due to early detection of plasma leakage and timely fluid replacement, women still develop per vaginal (PV) bleeding during dengue [15, 16]. Due to the limited data on the frequency of PV bleeding in women with dengue and associated complications and disease outcomes, we proceeded to characterize the frequency and disease severity in a large cohort of female patients with dengue in Sri Lanka.

## Methods

### Patients with acute dengue

288, female patients aged over 18 years of age, who were admitted to the National Institute of Infectious Disease, for management of a probable dengue infection were recruited following informed written consent from November 2021 to October 2024. An acute dengue infection was confirmed in them by either a rapid dengue NS1 antigen test (SD Biosensor, South Korea) or a quantitative PCR assay [17]. All patients were followed up from the time of admission to discharge from hospital and clinical and laboratory features including dengue warning signs and bleeding manifestations were recorded daily. The liver transaminases and C-reactive protein levels were recorded at the time of admission to hospital.

Abdominal ultrasound scans were done to assess the presence of fluid leakage in the form of pleural effusions or ascites in all patients at the time of admission and twice a day after that, throughout the period while they were in hospital, if they did not have features of plasma leakage in the initial scan. The treatment received including administration of fluid boluses, colloids and blood products was also recorded from their clinical records. The duration of hospital stay was recorded from time to admission to discharge.

Disease severity was classified according to the 2011 World Health Organization (WHO) dengue diagnostic criteria for dengue based on the disease progression while they were in hospital [18]. Accordingly, patients with a rise in haematocrit >20% of the baseline together with thrombocytopenia with accompanying bleeding manifestations, or those who had ultrasound scan evidence of plasma leakage were classified as having DHF. Shock was defined as the presence of a narrowing pulse pressure of 20 mm Hg in patients with DHF. According to the above criteria, 77 were classified as having DHF, and 211 patients as DF.

### Ethics approval

Ethical approval was obtained from the Ethics Review Committee of the Faculty of Medical Sciences, University of Sri Jayewardenepura (ethics application number:58/19). All patients gave informed written consent to participate in the study.

### Statistical analysis

Statistical analysis was carried out using GraphPad PRISM version 10.3.1. The degree of associations between disease severity and clinical and demographic characteristics, was expressed as the odds ratio (OR), which was obtained from standard contingency table analysis by Haldane’s modification of Woolf’s method. Chi Square tests or the Fisher’s exact test was used to determine the p value.

## Results

Of the 288 females recruited, 28 (9.72%) reported to have per vaginal bleeding, while 260 (90.27%) did not develop any bleeding manifestations. 77 (26.73%) of them were classified as DHF. 12/28 (42.85%) of those with PV bleeding had DHF, while 16, were classified as having DF, due to the absence of fluid leakage in repeated abdominal ultrasound scans. Therefore, women who developed PV bleeding were more likely to have developed DHF (OR 2.2, 95% CI 0.98 to 5.1, p=0.06). The clinical and laboratory features of the females with and without PV bleeding is shown in table 1. The women who had bleeding PV did not have any other overt bleeding manifestation.

**Table 1:** Clinical and laboratory features of those who developed PV bleeding vs those who had no bleeding manifestations.

| Clinical feature | Females who no had PV bleeding<br>N= 260 (%) | Females who had PV bleeding<br>N=28 (%) |
| --- | --- | --- |
| Abdominal pain | 90 (34.61) | 15 (53.57) |
| Vomiting | 104 (40) | 16 (57.14) |
| Diarrhoea | 76 (29.23) | 18 (64.28) |
| Gall bladder oedema | 104 (40) | 16 (57.14) |
| Any evidence of fluid leakage | 64 (24.61) | 11 (39.28) |
| Pericholecystic fluid | 55 (21.15) | 11 (39.28) |
| Free Fluid in hepatorenal pouch | 45 (17.30) | 6 (21.42) |
| PCV increase from baseline |  |  |
| 5-10% | 102 (39.23) | 9 (32.14) |
| 10-20% | 82 (31.53) | 9 (32.14) |
| >20% | 25 (9.61) | 5 (17.85) |
| Platelet counts |  |  |
| >100,000 | 66 (25.38) | 5 (17.85) |
| 50,000-100,000 | 105 (40.38) | 9 (32.14) |
| 20,000-49,999 | 65 (25) | 10 (35.71) |
| <20,000 | 25 (9.61) | 4 (14.28) |
| AST |  |  |
| <40 | 40 (15.38) | 4 (14.28) |
| 40-400 | 116 (44.61) | 11 (39.28) |
| >400 | 15 (5.76) | 0 (0) |
| ALT |  |  |
| <40 | 81 (31.15) | 9 (32.14) |
| 40-400 | 131 (50.38) | 13 (46.42) |
| >400 | 12 (4.61) | 0 (0) |
| CRP |  |  |
| <6 | 42 (16.15) | 1 (3.57) |
| 6 to 20 | 84 (32.30) | 11 (39.28) |
| 20-50 | 72 (27.69) | 5 (17.85) |
| >50 | 25 (9.61) | 2 (7.14) |
| Leucopenia | 61 (23.46) | 8 (28.57) |
| NS1 antigen positivity | 159/224 (70.98) | 18/22 (81.81) |

The duration of illness at the time of presentation for the patients was a median of 4 (IQR 3 to 4 Days). There was no significant difference (p= 0.64) in the day of presentation to hospital in those with bleeding PV (median 4, IQR 3 to 4) compared to those who had no bleeding (median 4, IQR 3 to 4). Those with PV bleeding were more likely to have abdominal pain (OR=2.17, 95% CI = 0.99 to 4.69, p = 0.06), vomiting (OR= 2.0, 95% CI= 0.89 to 4.44, p= 0.10), diarhoea (OR= 4.35, 95% CI = 1.908 to 9.610, p= 0.0004) or evidence of any fluid leakage (OR= 1.98, 95% CI = 0.91 to 4.5, p = 0.11) (table 10. They were also more likely to have a PCV rise of >20% from baseline (OR= 1.87, 95% CI = 0.72 to 5.34, p = 0.22), compared to those who did not have bleeding. Although there were no differences in the liver transaminase and C-reactive protein levels in women who developed bleeding PV, vs those who did not, those who had bleeding PV were more likely (OR= 1.92, 95% CI = 0.91 to 4.04, p – 0.14) to have a platelet count of <50,000 cells/mm3 (50%) than those who had no PV bleeding (34.6%). Interestingly, those who had PV bleeding were also more likely to have a positive NS1 antigen test (OR=1.84, 95% CI=0.59 to 5.17, p-0.33).

### Complications and treatment received by those with bleeding PV compared to others

As per standard care of this hospital, women who develop PV bleeding are given norethisterone 5mg three time a day and tranexamic acid 1g three times a day, until bleeding stops. The differences in other standard of care and the treatment received in these two groups is shown in table 2. Although not significant, those who had PV bleeding were more likely to have been given IV fluids, blood transfusions, dextran and IV antibiotics.

**Table 2:** The treatment received by female patients with dengue who either had PV bleeding or did not have any bleeding manifestations.

| Clinical feature | Females who had<br>no PV bleeding<br>N= 260 (%) | Females who had PV<br>bleeding<br>N=28 (%) | Odds ratio, 95%<br>CI and p value |
| --- | --- | --- | --- |
| IV fluid given | 172 (66.17%) | 21 (75%) | 1.53, 0.62 to 4.01,<br>p= 0.40 |
| Blood given | 11 (4.23%) | 3 (10.71%) | 2.71, 0.76 to 9.99,<br>p= 0.14 |
| Dextran given | 7 (2.69%) | 2 (7.12%) | 2.78, 0.55 to<br>13.79, p= 0.21 |
| Duration of hospital stay<br>(median, IQR) | 3 (3 to 4) | 4 (3 to 4.74) | p=0.43 |
| IV antibiotics given | 15 (5.76%) | 3 (10.71%) | 1.96, 0.56 to 6.43,<br>p= 0.39 |

## Discussion

In this study we sought to identify the frequency, risk factors and clinical outcomes in females with dengue, who developed PV bleeding, compared to the women who had no bleeding manifestations. We found that women who developed PV bleeding were more likely to have dengue warning signs such as abdominal pain, persistent vomiting and plasma leakage. They were also more likely to have a rise in PCV of ≥20% from baseline and platelet counts of ≤50,000 cells/mm^3^. Interestingly, although not considered a dengue warning sign, those who developed PV bleeding were significantly more likely to have diarrhea. Although not significant, they were more likely to require interventions such as blood transfusions, administration of dextran and intravenous antibiotics. The women in our cohort who developed PV bleeding, did not have worse disease outcomes, possibly as the standard of care in this hospital is to administer norethisterone and tranexamic acid as soon as PV bleeding is detected. This practice could have led to women with PV bleeding in this hospital, not having worse outcomes as reported elsewhere.

Although PV bleeding is a recognized complication of patients with dengue, the frequency and associated clinical outcomes have not been described [15, 16, 19]. PV bleeding in dengue may manifest as excessive bleeding in women who were menstruating at the time of illness, or intermenstrual bleeding [19]. Female gender was found to be associated with a higher risk of clinically significant bleeding, with 44.5% of those with clinically significant bleeding reporting menorrhagia [20]. Therefore, the higher risk of severe disease in females reported in many studies, could be due to PV bleeding [6, 13], although the reasons for classifying patients as having severe dengue is not explained. There is lack of data of characterizing the extent of PV bleeding and its possible contribution to disease outcomes in patients. For instance, a systematic review and metanalysis, which had only four studies recording PV bleeding, showed that bleeding from the gastrointestinal tract, pleural effusions, ascites and gall bladder wall thickening were significantly associated with development of severe dengue, in patients with dengue warning signs [21]. Although PV bleeding was shown to associate with a moderate risk of severe dengue (odds ratio 6.62), this was not statistically significant [21]. Therefore, more data is needed to understand the higher risk of severe dengue and mortality in females, with possible contribution of unrecognized PV bleeding.

Bleeding per vagina in acute dengue is likely to be due to the increased bleeding diathesis, a characteristic feature in dengue. Even though bleeding per vagina usually does not cause significant volume depletion in a healthy person, it is likely to contribute to intravascular volume depletion in a patient with plasma leakage. On the other hand, worsened bleeding diathesis in DHF especially when there is significant volume depletion will aggravate bleeding per vagina leading to a vicious cycle. Therefore, early identification of bleeding per vagina in dengue patients, initiation of treatment to reduce bleeding and prevention of volume depletion, is of paramount importance in reducing morbidity and mortality of dengue.

Dengue outbreaks have occurred in Sri Lanka for over three decades, with the number of cases rising each year [4]. Despite the increase in number of cases, Sri Lanka has managed to gradually reduce the CFRs from approximately 1% in 2009 to 0.05% by 2024 [4, 14]. Although a significant reduction in CFRs has been achieved over the years by public health education to seek medical care early, physician training programs for early detection of complications and meticulous and timely fluid management, 22 individuals succumbed to their illness in 2024. Since 19/23 of those who passed away are females, it would be important to understand the reasons for gender discrepancies in mortality rates in Sri Lanka and the rest of the world, especially in women in the reproductive age groups. Although gender differences in mortality have been observed with lower mortality rates among females with sepsis and COVID-19 [22, 23], females appear to have higher morbidity and mortality due to HIV [24]. Therefore, it would be important to conduct regular mortality reviews and study epidemiological and immunological mechanisms that lead to higher mortality in females with dengue, for understanding the causes of fatal dengue.

## Data Availability

The data is available within the manuscript and tables.

## Acknowledgments

We are grateful to the NIH, USA (grant number 5U01AI151788-02) for funding.

